# Allostatic (over)Load Measurement: Workflow and repository

**DOI:** 10.1101/2025.07.31.25332519

**Authors:** Shawna Beese, Jason Cross, David Rice, Trey L. DeJong

## Abstract

Researchers have long studied allostatic (over)load as an estimated measure of individual cumulative stress over a lifetime. Often called the overall ‘wear and tear’ from social and environmental stressors, allostatic (over)load shows promise as a practical indicator of general health trends in community settings. This data processing workflow aims to document our overall approach and reasoning when calculating allostatic (over)load for data analysis and knowledge sharing. The included repository features an R script for generating datasets using this workflow from the following data sources:

- *All of Us* Research Program data repository
- Health and Retirement Study (HRS)
- National Health and Nutrition Examination Survey (NHANES)

Our allostatic (over)load measurement process, along with the linked repository, provides a reproducible workflow to process secondary data and offers insights into protocol-driven measurement practices in community environments.

## BACKGROUND

Researchers have utilized allostatic (over)load as a conceptual model for the maladaptive biological processes that develop when our fight-or-flight acute stress response is activated as a chronic condition (Juster & Misiak, 2023; McEwen, 2000). For over two decades, researchers across various disciplines have also employed allostatic (over)load as an inferred measure of individual-level cumulative stress burden throughout the life course (Beckie, 2012). Often referred to as the overall ‘wear and tear’ from socio-environmental stressors (Solís et al., 2016; McEwen & Seeman, 1999), high allostatic load levels are a preclinical indicator of future chronic disease (e.g. cardiovascular disease, immune disorders, diabetes, anxiety, asthma) development (McEwen & Lasley, 2002).

Since the original 10 biomarkers were first reported in the literature (Seeman et al., 1997), several biomarker panels have been proposed as a potential standardized method for calculating allostatic (over)load (Beese et al., 2022; Duong et al., 2017). A comprehensive meta-analysis identifies a battery of five biomarkers that are most strongly associated with early indicators of generalized multi-system dysfunction resulting from the overburden of stress response among individuals in mid-life and beyond (McCrory et al., 2023). The five biomarkers index identified was based on the strength of association and biomarker availability. Our team acknowledges that there is a demand for more statistically nuanced methods for measuring allostatic (over)load in research, especially when studying pathway-specific physiological processes (Carbone et al., 2022; Liu et al., 2021). However, as a research team interested in allostatic (over)load as a generalized outcome variable that predicts overall health and mortality risk, we find that the five-biomarker index articulated in McCrory et al. has practical applications for representing the cumulative individual-level multi-system stress response (2023) for both data science and community research.

## METHOD

Initially, our data processing approach began with an evaluation of the *All of Us* database for the study of perceived stress and three different methods of calculating allostatic (over)load (Beese et al., 2024). Our refined data processing method for calculating allostatic (over)load, outlined in this document, initially utilized NIH *All of Us* Research Program (Table 1). This procedure resulted in a sample of 27,922 unique participants (Supplementary Table 1) from the NIH *All of Us* respondents with version 8 (V.8) controlled tier data. The Participant Identification Number (PIN) was used to merge fragmented data frames into a single, complete dataset. According to the *All of Us* data documentation, two standard operating procedures were used for collecting physical measurements (*All of Us* Research Program, n.d).

**Table 1.** Summary of Allostatic (over)Load Variable Calculation using NIH *All of Us* V.8.

| Allostatic (over)load biomarkers | Variable name in *All of Us* | *All of Us* repository location |
| --- | --- | --- |
| ***C-reactive protein (CRP)*** | C reactive protein [Mass/volume] in Serum | Electronic Health Record (EHR) |
| **-or-** | Plasma | ⇒ Labs & Measurements |
| ***High Sensitivity C-reactive protein (HS-CRP)*** | -or- |  |
|  | C reactive protein [Mass/volume] in Serum |  |
|  | Plasma by High sensitivity method |  |
| ***Heart Rate\**** | Computed heart rate, mean heart rate of 2<sup>nd</sup> and 3<sup>rd</sup> measures | Measurements & Wearables |
|  |  | ⇒ Physical Measurements |
| ***High Density Lipoprotein (HDL)*** | Cholesterol in HDL [Mass/volume] in Serum or Plasma | Electronic Health Record (EHR) |
|  |  | ⇒ Labs & Measurements |
| ***Glycated Hemoglobin (A1C)*** | Hemoglobin A1c/Hemoglobin total in Blood | Electronic Health Record (EHR) |
|  |  | ⇒ Labs & Measurements |
| ***Waist-to-Height Ratio (WHtR)\**** | Computed waist circumference, mean of closest two measures | Measurements & Wearables |
|  |  | ⇒ Physical Measurements |
|  | -and- |  |
|  | Height |  |

**Note:** \*For all unique identifiers in NIH *All of Us*, physical measurement values are present; the completeness of laboratory biomarker data limited the total sample size to 27,922.

### Summary of Physical Measurements Calculation

a. Heart Rate (*All of Us* Research Program, 2017, June)
  ∘ Mean of the 2nd and 3rd measures in the case of repeated measures.
b. Waist-to-Height Ratio (WHtR)
  ∘ This variable was calculated by first determining the calculated waist circumference (*All of Us* Research Program, 2017, October), and then dividing it by the calculated height (*All of Us* Research Program, 2017b, June).
  ∘ The mean of the two closest measures is used in the case of repeated measures.

### Summary of Biomarkers Data Cleaning and Processing

The following data processing approach (Steps 1-4) was used for the two physical measurement biomarkers, and the three laboratory biomarkers: a) High-density Lipoprotein (HDL), b) Glycated Hemoglobin (A1C), and c) C-reactive protein (CRP).

#### 1. Ensure only comparable lab measurements were used

a. Of all the different lab tests that could be used to represent each of the laboratory biomarkers, we used ONLY the most common test with established acceptance in the literature (e.g., serum or plasma test on HDL; Hemoglobin A1c/Hemoglobin total in blood).
b. The one exception to the above was that both serum or plasma CRP and serum or plasma high-sensitivity C-reactive protein (HS-CRP) are established in the literature. We cleaned the data for participants who had either-but kept the values sorted with separate CRP and HS-CRP columns. If a participant had values from both, we only used their CRP in the allostatic (over)load calculation. This procedure was used to ensure no duplicate weighing of CRP values for any given participant. CRP is preferred over HS-CRP because it also has a clinical threshold established in the literature (Table 2). The CRP clinical threshold cannot be used for HS-CRP.

**Table 2.** Clinical Thresholds for Potential Sensitivity Analysis.

| **Allostatic (over)load biomarkers and clinical threshold** | **Citation** |
| --- | --- |
| **Pulse or Heart rate (HR): $\geq 90$ beats/min** | **Parker et al., 2022<br>⇒ Borell et al., 2020<br>⇒ Howard & Sparks, 2016<br>⇒ Levine & Crimmins, 2014 |
| **High-Density Lipoprotein (HDL): $< 40$mg/dl** | **Parker et al., 2022<br>⇒ Akinyemiju et al., 2020<br>⇒ Borell et al., 2020<br>⇒ Howard & Sparks, 2016<br>⇒ Levine & Crimmins, 2014 |
| **C-Reactive Protein (CRP): $\geq 3.0$mg/L** | **Parker et al., 2022<br>⇒ Akinyemiju et al., 2020<br>⇒ Borell et al., 2020<br>⇒ Howard & Sparks, 2016<br>⇒ Levine & Crimmins, 2014 |
| **Glycated Hemoglobin (A1C): $\geq 6.4\%$** | **Parker et al., 2022<br>⇒ Borell et al., 2020<br>⇒ Howard & Sparks, 2016<br>⇒ Levine & Crimmins, 2014 |
| **Waist-to-Height Ration (WHtR): $> 0.5$** | Browning et al., 2010 |
| **\* Waist circumference (WC): $\geq 102$ cm (male)/ $\geq 88$ cm (female)** | Seo et al., 2017 |

**Note:** \*In addition to waist-to-height ratio as the preferred anthropometric measure (McCrory et al., 2023), we additionally provide a clinical threshold for waist circumference. \*\*Parker et al., 2022, published these meta-analysis clinical cutoffs.

#### 2. Determine if there were repeated measures

a. Some individual respondents had multiple observations recorded on different dates (repeated measures). To establish a single metric for each individual, the mean was calculated over the recorded metrics for an individual within one year from the most recent measurement date.
b. Additionally, the observation was removed if only NA values were recorded for the individual.

#### 3. Removal of extreme outlier values

a. We set the initial outlier cutoffs for each biomarker as four standard deviations from the mean.
b. Cutoff ranges were further assessed for clinical relevance in the literature. Coherence between statistical and clinical cutoffs was confirmed.

#### 4. Calculate the allostatic (over)load risk category

a. The top quartile CRP, heart rate, A1C, and WHtR were coded as 1=high risk.
b. The bottom quartile of HDL was coded 1=high risk, as HDL is a cardio-protective biomarker.
c. All observations determined to be in the lower 75%-tiles of risk were coded 0=normal risk.

Each participant is assigned a cumulative score of 0-5 (with 0 being the lowest and 5 being the highest calculated risk) based on the sum of their high-risk scores across all five biomarkers. Although we acknowledge that statistical modeling power is lost by dichotomizing each biomarker into high-normal risk, this is the most utilized method and the method presented in the McCrory et al., 2023 consensus article. All biomarkers must be present to calculate a respondent’s overall score.

### Recommendations for Clinical Cutoffs

For future studies, we plan to utilize additional clinical cutoffs established in the literature to determine high-risk and normal-risk categories, and report findings from both methods as part of a sensitivity analysis. Below are our recommended clinical thresholds (Table 3). To calculate allostatic (over)load, use the same schema as above (step 4).

**Table 3.** Summary of Allostatic (over)Load Variable Calculation using Health and Retirement Survey (HRS)

|  | **Biomarker**<br>**Data** | **HRS CORE**<br>**Data** |
| --- | --- | --- |
| **2016** | HDL | Pulse (heart rate) |
|  | A1C | Waist-to-height ratio (WHtR) |
|  | CRP | Demographics (age, sex at birth, race, ethnicity) |
| **2014** | HDL | Pulse (heart rate) |
|  | A1C | Waist-to-height ratio (WHtR) |
|  | CRP | Demographics (age, sex at birth, race, ethnicity) |
| **2012** | HDL | Pulse (heart rate) |
|  | A1C | Waist-to-height ratio (WHtR) |
|  | CRP | Demographics (age, sex at birth, race, ethnicity) |
| **2010** | HDL | Pulse (heart rate) |
|  | A1C | Waist-to-height ratio (WHtR) |
|  | CRP | Demographics (age, sex at birth, race, ethnicity) |
| **2008** | HDL | Pulse (heart rate) |
|  | A1C | Waist-to-height ratio (WHtR) |
|  | CRP | Demographics (age, sex at birth, race, ethnicity) |
| **2006** | HDL | Pulse (heart rate) |
|  | A1C | Waist-to-height ratio (WHtR) |
|  | CRP | Demographics (age, sex at birth, race, ethnicity) |

**Note:** HDL= High-density lipoprotein; A1C= Glycated hemoglobin; CRP= C-reactive protein.

### HRS and NHANES Datasets

Allostatic (over)load is calculated for the Health and Retirement Study (HRS; Table 3; Supplementary Table 2) and National Health and Nutrition Examination Survey (NHANES; Table 4; Supplementary Table 3) with the same approach as described for the NIH *All of Us*, with minor adaptations.

**Table 4.**
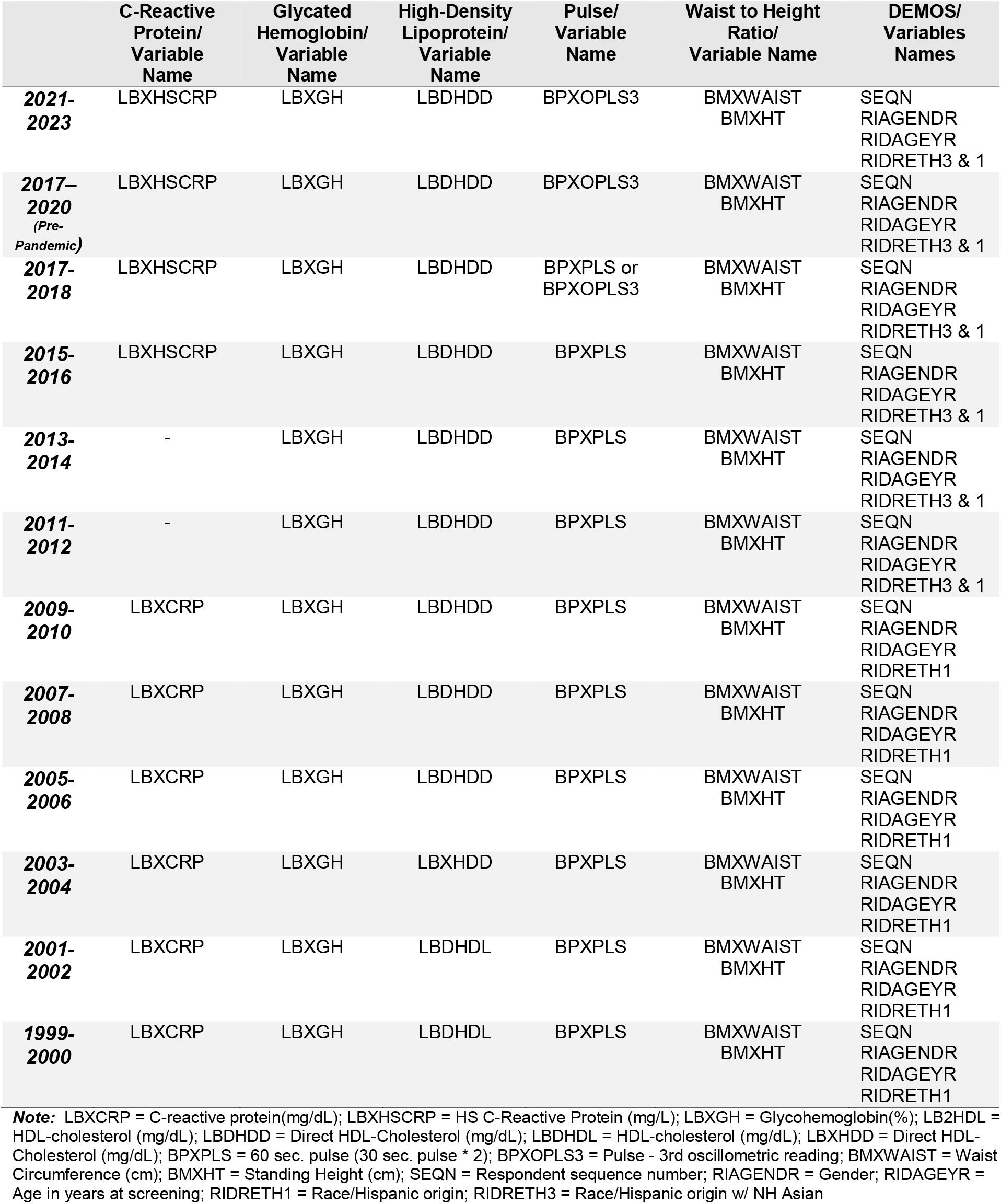
Summary of Allostatic (over)Load Variable Calculation using National Health and Nutrition Examination Survey (NHANES)

1. Pulse (heart rate) as a single data point is used, as opposed to a mean of repeated measures for Allostatic (over)Load calculation with the Health and Retirement Study (HRS) and National Health and Nutrition Examination Survey (NHANES) as a data source.
2. Only C-reactive protein (CRP) is used with the Health and Retirement Study (HRS). Both CRP and high-sensitivity C-reactive protein (HS-CRP), depending on availability, are used with the National Health and Nutrition Examination Survey (NHANES) as a data source.

### Community Collected Biomarker Measurements

When collecting allostatic (over)load metrics as primary sourced data in community settings, we recommend contracting with an appropriately credentialed lab for the collection of:

- High-density Lipoprotein (HDL)
- Glycated Hemoglobin (A1C)
- C-reactive protein (CRP)

Physical measurements will likely be collected independently at the community site by internal staff. We present the following PhenX protocols, as the PhenX Toolkit aims to provide widely accepted, standardized measurement procedures across various research domains (Cox et al., 2021).

a. Resting Heart Rate (PhenX Toolkit. n.d.-a)
b. Waist-to-Height Ratio (WHtR)
  ∘ This variable will be computed by first calculating the waist circumference (PhenX Toolkit, n.d.-b), and then dividing it by the calculated standing height (PhenX Toolkit, n.d.-c).

## Limitations and Future Research Implications

A limitation of this approach is that it is grounded in research that focused explicitly on mid-life and beyond. Future synthesis (e.g., meta-analysis) research into the temporal variations of allostatic (over)load across the life course is still warranted.

A perennial critique of allostatic (over)load calculation is that it is dichotomized into a high-risk/low-risk schema. A dichotomized schema has been used for decades and has been justified in the literature (McCrory et al., page 6, para 3, 2023). However, we do recommend that future research use sensitivity analyses with established clinical thresholds. Also, if future researchers develop other categorical sub-classifications (e.g., low 0-2; moderate 3; high 4-5), sensitivity analysis with the classic method is also recommended, as this would establish a comparative context to practices already established in the literature.

## CONCLUSION

The data processing workflow, along with our linked repository, are aimed to provide a transparent and reproducible method by which to calculate allostatic (over)load. Additionally, we are sharing our process as a resource to encourage discourse with other researchers and data scientists who are also studying allostatic (over)load as a generalized variable to infer preclinical maladaptive responses to stressors that often precede the development of future chronic diseases.

## Author Contributions

Conceptualization, S.B.; methodology, D.R., J.C., T.L.D., and S.B.; data curation, D.R., J.C., T.L.D., and S.B.; writing—original draft preparation, S.B.; writing—review and editing, D.R., J.C., T.L.D., and S.B; GitHub repository management, J.C.; project administration, S.B. All authors have read and agreed to the published version of the manuscript.

## Data Availability Statement

This study utilized third-party publicly accessible data made available at: National Health and Nutrition Examination Survey (NHANES) at https://wwwn.cdc.gov/nchs/nhanes/

This study utilized third-party data made available under a license for which the authors do not have permission to share. Requests to access the data should be directed to: *All of Us* Research Program data repository at https://workbench.researchallofus.org/login; and Health and Retirement Study (HRS) at https://hrs.isr.umich.edu/data-products

R Markdowns and documentation supporting this study are openly available from CHORDS Lab GitHub at: https://github.com/CHORDSLab/Allostatic-over-Load-Repository

## ACKNOWLEDGMENT

The authors would like to extend deep gratitude to the Data 424: Data Analytics Capstone Team 2 members— Mariah Bergquist, Maya Dietrich, William Ripplinger, and Owen Williams—for sharing their technical skills and expertise in the development of the National Health and Nutrition Examination Survey (NHANES) dataset. We also want to thank lab member Kristen Barta for her final proofreading of this manuscript.

We want to express our appreciation to all participants in the NIH *All of Us* Research Program, the Health and Retirement Study (HRS), and the National Health and Nutrition Examination Survey (NHANES). Only through their contributions was this study made possible.

---

**Supplementary Table 1.** Demographic Characteristics for Allostatic (over)Load Sample from NIH *All of Us* V.8.

|  | *Female*<br>(N=17,339) | *Male*<br>(N=10,254) | *Overall*<br>(N=27,922) |
| --- | --- | --- | --- |
| **Race** |  |  |  |
| American Indian or Alaska Native | 188 (1.1%) | 158 (1.5%) | 352 (1.3%) |
| Asian | 292 (1.7%) | 169 (1.6%) | 468 (1.7%) |
| Black or African American | 3,415 (19.7%) | 1,485 (14.5%) | 4,974 (17.8%) |
| Middle Eastern or North American | 94 (0.5%) | 58 (0.6%) | 158 (0.6%) |
| More than one population | 816 (4.7%) | 392 (3.8%) | 1,227 (4.4%) |
| None indicated, none of these, skip | 2,475 (14.3%) | 1,263 (12.3%) | 3,789 (13.6%) |
| White | 10,059 (58%) | 6,729 (65.6%) | 16,954 (60.7%) |
| **Ethnicity** |  |  |  |
| Hispanic or Latino | 2,445 (14.1%) | 1,141 (11.1%) | 3,606 (12.9%) |
| Not Hispanic or Latino | 14,448 (83.3%) | 8,779 (85.6%) | 23,494 (84.1%) |
| Prefer not to answer, skip, none of these | 446 (2.6%) | 334 (3.3%) | 822 (2.9%) |
| **Age** |  |  |  |
| Mean (SD) | 55.3 (14.5) | 59.4 (14.0) | 56.9 (14.5) |
| Median [Min., Max.] | 56.6 [<18.0 ,99.7] | 61.0 [<18.0, 102.0] | 56.6 [<18.0 ,102.0] |

**Note:** Not all cross-tabs add up to 100% due to the masking of cells with n < 20; and n = 329 respondents identifying as a sex at birth other than Female or Male. All ages under 18 years are reported as <18 for the min.

**Supplementary Table 2.** Demographic Characteristics for Allostatic (over)Load Sample from Health and Retirement Survey (HRS)

|  | **2006**<br>**(N=18,469)** | **2008**<br>**(N=17,217)** | **2010**<br>**(N=22,034)** | **2012**<br>**(N=20,553)** | **2014**<br>**(N=18,747)** | **2016**<br>**(N=20,912)** | **Overall**<br>**(N=117,932)** |
| --- | --- | --- | --- | --- | --- | --- | --- |
| **Sex** |  |  |  |  |  |  |  |
| *Male* | 7,588 (41.1%) | 7,022 (40.8%) | 9,231 (41.9%) | 8,550 (41.6%) | 7,704 (41.1%) | 8,670 (41.5%) | 48,765 (41.4%) |
| *Female* | 10,881 (58.9%) | 10,195 (59.2%) | 12,803 (58.1%) | 12,003 (58.4%) | 11,043 (58.9%) | 12,242 (58.5%) | 69,167 (58.6%) |
| **Race** |  |  |  |  |  |  |  |
| *White/Caucasian* | 14,918 (80.8%) | 498 (2.9%) | 3,578 (16.2%) | 378 (1.8%) | 136 (0.7%) | 2,475 (11.8%) | 21,983 (18.6%) |
| *Black/African American* | 2,572 (13.9%) | 166 (1.0%) | 2,074 (9.4%) | 122 (0.6%) | 56 (0.3%) | 1,321 (6.3%) | 6,311 (5.4%) |
| *Other (includes Native/Asian/PI)* | 915 (5.0%) | 57 (0.3%) | 1,005 (4.6%) | 103 (0.5%) | 27 (0.1%) | 223 (1.1%) | 2,330 (2.0%) |
| *Don't Know/NA* | 52 (0.3%) | 3 (0.0%) | 48 (0.2%) | 8 (0.0%) | 1 (0.0%) | 31 (0.1%) | 143 (0.1%) |
| *Refused* | 12 (0.1%) | 0 (0%) | 14 (0.1%) | 2 (0.0%) | 0 (0%) | 6 (0.0%) | 34 (0.0%) |
| *Missing* | 0 (0%) | 16,493 (95.8%) | 15,315 (69.5%) | 19,940 (97.0%) | 18,527 (98.8%) | 16,856 (80.6%) | 87,131 (73.9%) |
| **Ethnicity** |  |  |  |  |  |  |  |
| *Mexican American/Chicano* | 1,044 (5.7%) | 58 (0.3%) | 863 (3.9%) | 80 (0.4%) | 15 (0.1%) | 504 (2.4%) | 2,564 (2.2%) |
| *Other Latino/a (includes Puerto Rican, Cuban)* | 658 (3.6%) | 45 (0.3%) | 616 (2.8%) | 68 (0.3%) | 24 (0.1%) | 484 (2.3%) | 1,895 (1.6%) |
| *Refused* | 1 (0.0%) | 0 (0%) | 1 (0.0%) | 0 (0%) | 0 (0%) | 2 (0.0%) | 4 (0.0%) |
| *Missing* | 16766 (90.8%) | 17,114 (99.4%) | 20,554 (93.3%) | 20,405 (99.3%) | 18,708 (99.8%) | 19,922 (95.3%) | 113,469 (96.2%) |
| **Age** |  |  |  |  |  |  |  |
| *Mean (SD)* | 68.5 (11.1) | 69.7 (10.8) | 65.8 (12.0) | 67.2 (11.6) | 68.4 (11.3) | 65.8 (11.9) | 67.5 (11.6) |
| *Median [Min, Max]* | 68.0 [25.0, 105] | 69.0 [25.0, 108] | 65.0 [18.0, 109] | 66.0 [<18, 104] | 67.0 [<18, 105] | 64.0 [21.0, 107] | 67.0 [<18, 109] |

**Note:** Data source Health and Retirement Survey (HRS) core data (publicly reported) and biomarker data (requires access to sensitive data; permissions required). All ages <18 are reported as <18.

**Supplementary Table 3.**
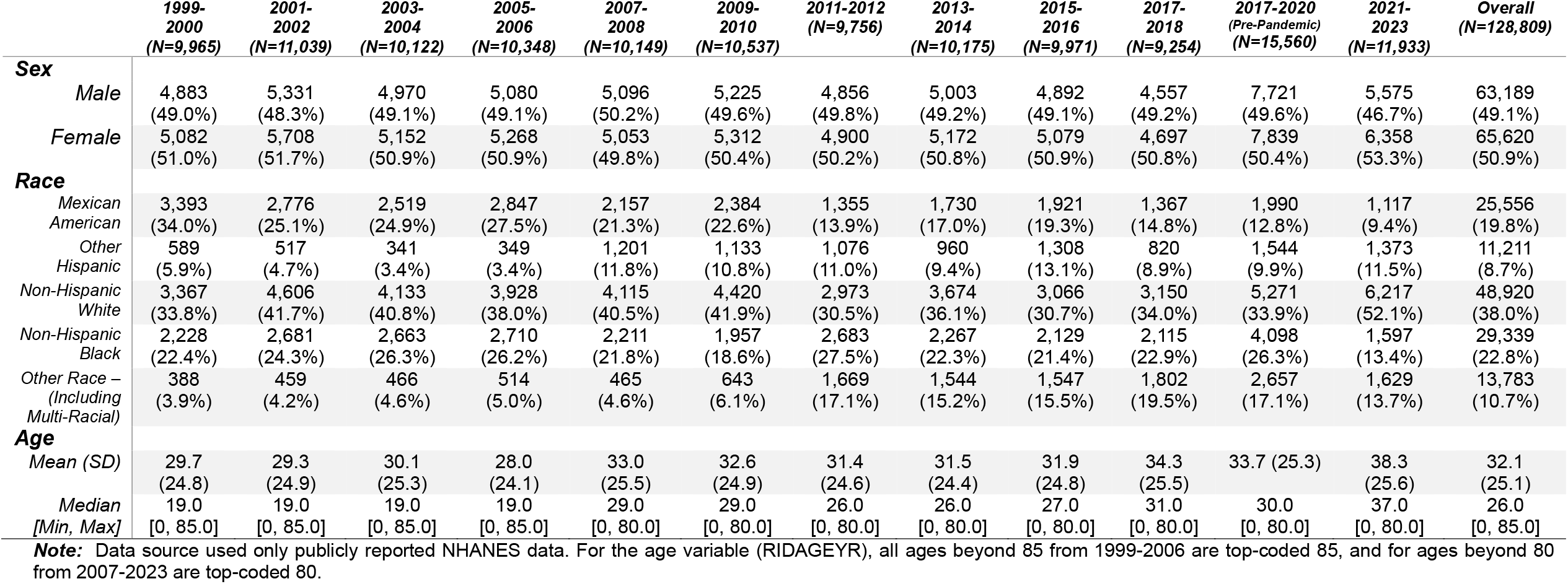
Demographic Characteristics for Allostatic (over)Load Sample from National Health and Nutrition Examination Survey (NHANES)

